# Corticosteroids and mortality in patients with severe Covid-19 who have autoantibodies

**DOI:** 10.1101/2021.03.19.21253005

**Authors:** Zhu Cui, Maxwell Roth, Yelena Averbukh, Andrei Assa, Azal Al Ani, William Southern, Morayma Reyes Gil, Shitij Arora

## Abstract

Auto-reactivity in COVID-19 is increasingly being recognized and may identify a group of patients with inflammation severe enough to result in loss of self-tolerance. Corticosteroids are potent anti-inflammatory agents and now the standard of care for patients with severe Covid-19 requiring oxygen support/mechanical ventilation. We studied the outcomes of COVID-19 patients who demonstrated clinically identifiable auto-reactivity and received corticosteroid treatment.

In this retrospective cohort study, we included 51 COVID-19 patients admitted between March 10, 2020 and May 2, 2020 who received corticosteroid treatment and also had serum sample in our institution bio-bank available for ANA and RF ELISA. Twelve patients (23.5%) had positive ANA or RF. Mortality rate among patients with positive autoantibodies was significantly higher than those without (9/12 or 75% versus 13/39 or 33.3%, p= 0.02). The high mortality rate in patients with auto-reactivity warrants further investigation and may be the subgroup where additional immunomodulation is effective.

## Introduction

Auto-reactivity in Coronavirus disease 2019 (COVID-19) is increasingly being recognized (1-3). Some of these autoantibodies may contribute to the development of severe disease or even long COVID symptoms that persist months after initial infection(3, 4). Autoantibodies are more commonly detected among patients with worse disease severity and higher CRP level (1, 2). They may identify a group of patients with inflammation severe enough to result in loss of self-tolerance(5). Corticosteroids are potent anti-inflammatory agents and now the standard of care for patients with severe Covid-19 requiring oxygen support/mechanical ventilation (6). We sought to study the outcomes of COVID-19 patients who demonstrated clinically identifiable auto-reactivity and received corticosteroid treatment.

## Methods

We identified adult patients hospitalized with severe COVID-19 between March 10, 2020 and May 2, 2020, who were initiated on corticosteroids within 48 hours of admission, and received at least two doses of treatment (0.5mg/kg/day prednisone equivalent or higher). Patients who passed away within 48 hours of admission were excluded (Figure 1). Serum samples collected during hospitalization were promptly aliquoted and stored at −80°C. Antinuclear antibody (ANA) and rheumatoid factor (RF) testing were conducted using EliA kits on the Phadia 250 instrument following the manufacturer’s instructions. Reporting units and cut-offs for determining positive result were pre-specified by the manufacturer (see detailed methods in the supplementary file).

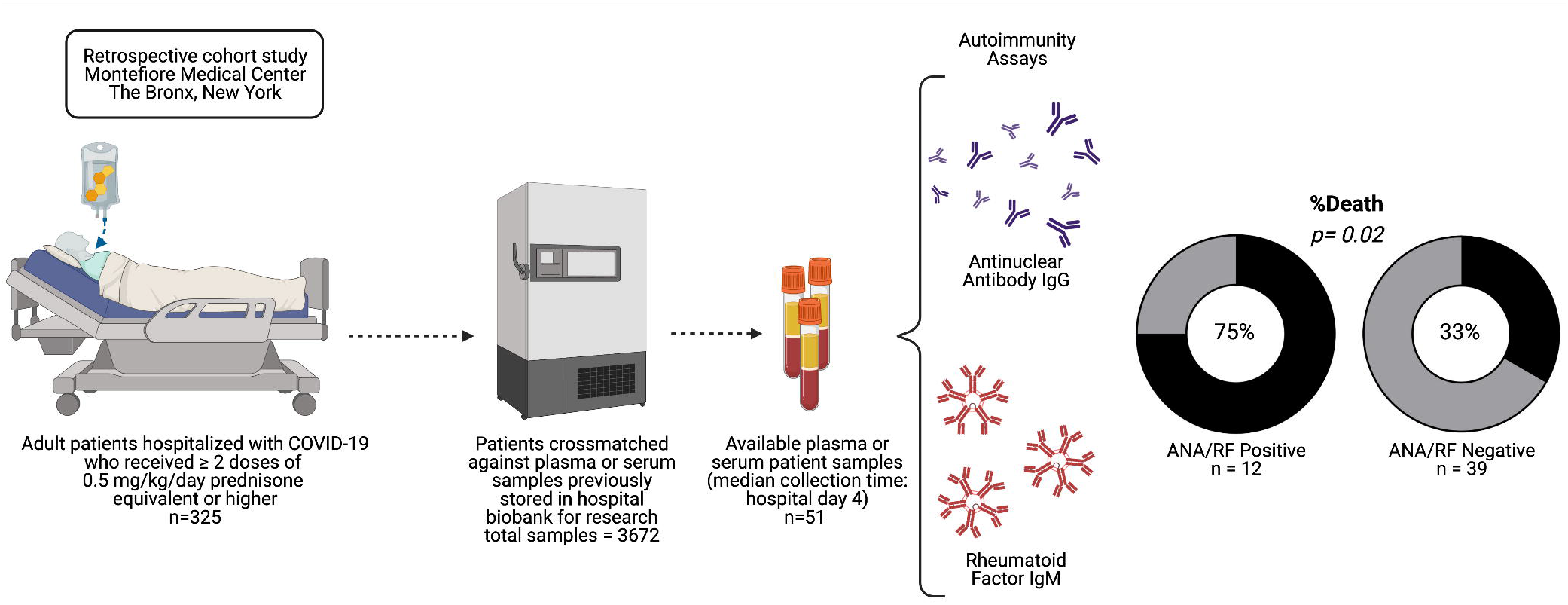

## Results

Serum samples were available for 51 patients, 12 (23.5%) were positive for either ANA or RF (six ANA positive, six RF positive), and 39 (76.5%) were negative for both (three patients with equivocal results were categorized as negative for analysis). Three patients with positive autoantibodies had previously diagnosed autoimmune disease and one of whom with known positive RF prior to COVID-19 infection. Patients with autoantibodies had higher Charlson Comorbidity Index, but similar admission lymphocyte counts, C-reactive protein, and D-dimer levels (Table 1). There were a total of 22 deaths (43%). Among the 12 patients with positive ANA/RF, nine died (75%), whereas out of the 39 patients who were ANA/RF negative, thirteen died (33%). The mortality difference was statistically significant (p=0.02). Rate of mechanical ventilation was also higher among autoantibody-positive patients but not statistically significant (41.7% versus 28.2%, p=0.48).

**Table 1.** **Patient baseline characteristics, clinical characteristics on admission, treatments received, and clinical outcomes**

|  |  |  |  |
| --- | --- | --- | --- |
|  | **ANA/RF Neg**<br>**(N=39)** | **ANA/RF Pos**<br>**(N= 12)** | **P**<br>**Value** |
| ***Baseline Characteristics*** |  |  |  |
| **Median Hospital Day of**<br>**Sample Collection (IQR)** | 4 (2 – 8) | 5 (1 – 7.5) | 0.98 |
| **Median Age (IQR)** | 64 (47 – 77) | 70 (59.5 – 83.5) | 0.17 |
| **Female (%)** | 19 (48.7) | 4 (33.3) | 0.51 |
| **Autoimmune Disease (%)** | 0 (0) | 3 (25.0) | 0.01 |
| **Prior ANA/RF (%)** |  |  | 0.23 |
| Positive | 0 (0) | 1 (8.3) |  |
| Negative | 2 (5.1) | 1 (8.3) |  |
| Unknown | 37 (94.9) | 10 (83.3) |  |
| **Ever Smoker (%)** | 1 (2.6) | 2 (16.7) | 0.13 |
| **Diabetes (%)** | 14 (35.9) | 5 (41.7) | 0.74 |
| **Median Charlson**<br>**Comorbidity Index (IQR)** | 3 (1 – 3) | 4 (2 – 7) | 0.04 |
| **Full Code (%)** | 33 (84.6) | 9 (75.0) | 0.42 |
| ***Clinical Characteristics On Admission*** |  |  |  |
| **Median Initial White Blood**<br>**Cell Count (IQR), k/mcL** | 8.4 (6.8 – 10.8) | 7.0 (5.4 – 12.6) | 0.50 |
| **Median Initial Neutrophil**<br>**Count (IQR), k/mcL** | 6.9 (5.6 – 9.1) | 6.2 (3.9 – 10.9) | 0.60 |
| **Median Initial Lymphocyte**<br>**Count (IQR), k/mcL** | 0.9 (0.6 – 1.2) | 0.75 (0.6 – 0.9) | 0.12 |
| **Median Initial Creatinine**<br>**(IQR), mg/dL** | 1.2 (0.8 – 1.9) | 1.3 (1.0 – 2.4) | 0.58 |
| **Median Initial Troponin**<br>**(IQR), ng/mL** | 0.01 (0.01 – 0.04) | 0.02 (0.01 – 0.06) | 0.4 |
| **Median Initial CRP (IQR),**<br>**mg/dL** | 15.8 (8.5 – 18.5) | 17.6 (15.0 – 33.7) | 0.17 |
| **Median Initial D-dimer**<br>**(IQR) ug/mL** | 1.8 (0.8 – 3.7) | 2.7 (1.1 – 5.9) | 0.14 |
| **Median Initial IL6 (IQR),**<br>**pg/mL** | 36.3 (10.4 – 126.6) | 145.0 (17.3 – 272.3) | 0.10 |
| ***Treatments Received*** |  |  |  |
| Hydroxychloroquine (%) | 31 (79.5) | 7 (58.3) | 0.29 |
| Sarilumab or Leronlimab (%) | 2 (5.1) | 4 (33.3) | 0.02 |
| Remdesivir (%) | 5 (12.8) | 1 (8.3) | 1 |
| ***Clinical Outcomes*** |  |  |  |
| **Highest O2 Requirement** |  |  |  |
| **(%):** |  |  |  |
| RA/NC | 12 (30.8) | 1 (8.3) | 0.15 |
| NRB | 11 (28.2) | 5 (41.7) | 0.48 |
| HFNC/CPAP/BiPAP | 5 (12.8) | 1 (8.3) | 1 |
| MV | 11 (28.2) | 5 (41.7) | 0.48 |
| **Median Hospital stay (IQR)** | 7 (5 -11) | 9 (4.5 -13) | 0.80 |
| **Death (%)** | 13 (33.3) | 9 (75.0) | 0.02 |

## Discussion

We report presence of autoantibodies among patients with severe COVID-19. Mortality rate was significantly higher among those with positive autoantibodies than those without. These findings are hypothesis generating. It may be possible that presence of autoantibodies reflects a state of hyper-inflammation where synergistic immunomodulation with corticosteroids and other targeted therapies (antagonists of interleukin-1 or interleukin-6) is more effective. Our study is limited by its small sample size, retrospective methodology, and types of autoimmune antibody tested. We were also unable to differentiate between pre-existing autoantibodies and those with ‘de-novo’ autoantibody production due to severe Covid-19. Nonetheless, the high mortality rate in patients with auto-reactivity warrants further investigation.

## Data Availability

The authors confirm that the data supporting the findings of this study are available within the article [and/or] its supplementary materials.

## Notes

There is no conflict of interest to declare.

### Competing Interest Statement

The authors have declared no competing interest.

### Funding Statement

No funding source to disclose.

### Author Declarations

This study was approved by the institutional review board of the Albert Einstein College of Medicine.

