## Supplemental Methods for "Corticosteroids and mortality in patients with severe Covid-19 who have autoantibodies"

*Antinuclear Antibody IgG (EliA Symphony) and Rheumatoid Factor IgM (Elia RF IgM) Assay*

Patient samples were thawed in a 37˚C water bath, diluted 1:100 in EliA Sample Diluent (PBS containing BSA, detergent and sodium azide), and then added to a 96 well MicroWell Dilution Plate. For the antinuclear antibody IgG (EliA Symphony) assay, the 96-well plate used was pre-coated with human recombinant U1RNP, SS-A/Ro, SS-B/La, Centromere B, Scl-70, Jo-1, and native purified Sm proteins. For the rheumatoid factor IgM (EliA RF IgM) assay, the 96-well plate used was pre-coated with aggregated rabbit IgG antigen. After a 30-minute incubation, a Washing Solution (Phosphate buffer containing detergent) was used to remove non-bound antibodies. Enzyme-labeled human IgG (EliA IgG conjugate) or IgM antibodies (EliA IgM conjugate) were then added and incubated for 28 minutes. Non-bound conjugate was washed away using the Washing Solution. Development Solution (0.01 % 4-Methylumbelliferyl-β-D-galactoside) was then applied to the wells for 39 minutes. After stopping the reaction using the Stop Solution (4% sodium carbonate), fluorescence in each well was measured at 447 nanometers. All incubations and measurements were performed at 37˚C and the instrument was calibrated to an international reference standard. Phadia 250 measures specific IgG or IgM concentrations in µg/l; the results were then converted to a ratio or IU/mL, respectively, using the lot-specific calibrators. A positive result is defined by the manufacturer as a ratio greater than 1.0 for the antinuclear antibody IgG assay and a value greater than 5.0 IU/mL for the rheumatoid factor IgM assay.
